# Intranasal lavage with hypochlorous acid safely reduces the symptoms in the ambulatory patient with COVID-19

**DOI:** 10.1101/2023.07.17.23292426

**Authors:** Monique Lisa Abner

## Abstract

**OBJECTIVE:** This study was designed to investigate intranasal lavage with a hypochlorous acid solution in the reduction of symptoms in the ambulatory COVID-19 patient.

**STUDY DESIGN:** Study approval granted by the Institutional Review Board of Reading Hospital (IRB 036-20), with informed consent obtained from all adult participants(age>18 years).

**SETTING:** All enrollees, taken from the same ambulatory testing facility, received nasopharyngeal swabs for COVID-19 testing by reverse transcription polymerase chain (RT-PCR) or the COVID-19 antigen specific test (Binax NOW, Abbott Lab)

**METHODS:** Convenience sampling methodology was utilized. Each enrollee was provided with the study devices which included a Nasaflo Neti Pot (NeilMed Pharmaceutical, Inc.), and the hypochlorous acid solution (Vashe Wound Solution, Urgo Medical North America, LLC). Participants were instructed to irrigate each nostril with 120 cc (four ounces) of the solution for ten consecutive days, and record the presence or absence of symptoms in a scripted diary log.

**RESULTS:** The study included 88 patients of which 74 (84.1%) completed the ten days of nasal lavage. All data analysis was conducted using SPSS version 25.0.

Chi square test of association found no significant difference related to gender, age group race, ethnicity, residence, or living arrangements (all p-values > 0.05). There were no statistical differences in any of the co-morbid conditions. Mild adverse reactions included burning, epistaxis, and oral metallic taste. No enrollees required mechanical ventilation. There were no deaths.

**CONCLUSION:** This study suggests the feasibility and safety of using intranasal lavage with a hypochlorous acid solution in relieving symptoms in the ambulatory Covid-19 patient.

## INTRODUCTION

The coronavirus, COVID-19, has a high affinity for the angiotensin-converting enzyme 2 receptor (ACE 2).^1^ There is a high expression of the ACE2 receptors in the goblet cells of the nasoepithelium which has been the basis for obtaining nasal and nasopharyngeal swabs for diagnostic purposes.^2,3^ Hypochlorous Acid is a weak acid that has been used clinically as a wound cleanser with favorable, non-cytotoxic activity against bacteria, virus, and some fungi. ^4,5,6^. The proposed use of a hypochlorous acid nasal lavage would be to inactivate the viral particles within the nasal tract. This study was designed to investigate the feasibility of intranasal lavage with hypochlorous acid as a therapeutic intervention in the reduction of symptoms of severe acute respiratory syndrome coronavirus 2 (SARS-CoV-2) in the ambulatory patient.

## METHODS

### Study Design and Participants

This study was approved by the Institutional Review Board of Reading Hospital (IRB 036-020) with informed consent obtained from all adult participants. Convenience sampling methodology was utilized for those who met the study criteria inclusive to all gender at ages18 years old and/or older. Excluded participants were those who expressed the desire to not use nasal lavage, women with known pregnancies, and children aged 17 years old and/or younger. The source of enrollees was generated from patients presenting to an ambulatory testing facility for nasopharyngeal swabs for COVID-19. Swab analysis was obtained by reverse transcription-polymerase chain reaction until the COVID-19 antigen specific test (Binax NOW, Abbott Lab) became available for use which allowed a more lenient process for including the Covid-19 positive patients. Each enrolled participant was given a Nasaflo Neti Pot (NeilMed Pharmaceuticals, Inc) with instructions to irrigate each nostril once a day for ten days with the provided hypochlorous solution Vashe Wound Solution (Urgo Medical North America, LLC). Each participant began the daily 120 cc (four -ounce) nasal lavage intervention within 72 hours of their testing. Each participant was given a scripted diary log in which they recorded daily the presence/absence of clinical symptoms such as elevated temperature, fatigue, headache, chills, nausea, and anosmia by noting “yes” or “no.“ After completing the ten days of nasal lavage, each log was returned to the principal investigator via mail. The participants remained quarantined for the recommended time period of 14 days, and adhered to social distancing, handwashing, and donning of face masks. Throughout the study, each participant was urged to not share their devices with any other individuals. After thirty days from the start of their nasal intervention, participants were telephoned for a follow-up review.

## RESULTS

A total of 88 patients were enrolled in the study, of which 74 (84.1%) completed the ten days of intranasal lavage. The reasons for the 14 patients withdrawing included the following: complaints of nasal burning, coughing, and inexperience with the nasal lavage (n=8); discouragement by another family member, being too busy to comply (n=2); and, negative Covid test and no specific reason given (n=4). No patients required hospital admission for mechanical ventilation. Due to the smaller than expected sample size, responders were grouped into 2 groups, those that felt their symptoms had stayed the same or worsened, and those who felt that their symptoms had improved. One patient felt that symptoms had both improved and worsened, and was placed into the worsened cohort. Small samples within certain age groups allowed for clustering of ages into 2 groups youngest to age 49, 50 years of age and older. Two patients did not respond to the question of race, and three patients did not respond to the question of living arrangements; for those items they were left out of the analysis.

All data analysis for this research was conducted using SPSS version 25.0. Since all variables in this analysis were categorical comparisons on patient perceived condition (worsened/stayed the same versus improvement) as the dependent variable, data were compared against comorbidities and demographic variables using chi-square test of association. An a priori p-value required for significance was set at 0.05 (p<0.05). Due to the exploratory nature of this research, there were no corrections applied to the p-value’s due to multiple comparisons.

Within the 74 patients that completed the course of treatment, 56.8% of the sample were females, age was evenly distributed among 6 age groups. 69.4% of the sample was Caucasian with the majority of those evaluating themselves as non-Hispanic. 90.5% of the sample resided in a single-family home with 85.9% of the sample indicating they lived with a spouse or significant other. Results of the demographic analysis can be found in Table 1.

**Table 1:** Demographic characteristics of completing patients (n=74).

| Variable | Category | Count | Percent |
| --- | --- | --- | --- |
| Gender | Female | 42 | 56.8 |
|  | Male | 32 | 43.2 |
| Age group | Youngest – 30 | 15 | 20.3 |
|  | 31-39 | 15 | 20.3 |
|  | 40-49 | 15 | 20.3 |
|  | 50-60 | 14 | 18.9 |
|  | 61-70 | 11 | 14.9 |
|  | 71-80 | 4 | 5.4 |
| Race* | African American | 3 | 4.2 |
|  | Asian | 2 | 2.8 |
|  | Caucasian | 50 | 69.4 |
|  | Multiple | 4 | 5.6 |
|  | Other | 13 | 18.1 |
| Ethnicity | Non-Hispanic | 46 | 62.2 |
|  | Hispanic | 28 | 37.8 |
| Residence | Apartment | 2 | 2.7 |
|  | Condo | 2 | 2.7 |
|  | Multigenerational | 3 | 4.1 |
|  | Single Family Home | 67 | 90.5 |
| Lives with** | Alone | 4 | 5.6 |
|  | Spouse/Significant other | 61 | 85.9 |
|  | Parent | 4 | 5.6 |
|  | Other | 2 | 2.8 |

\*There were two non-responders. \*\*Three non-responders

Chi-square test of association found no significant difference in patient condition related to patient gender, age group race, ethnicity, residence or living arrangements, (all p-values were greater than 0.05). A significant association (difference) was found for the recoded age variable, finding that 65.2% of the youngest through age 49 cohort reported improved condition as compared to only 34.8% of those in the 50 through oldest years of age cohort (p = 0.028). These results can be found in Table 2.

**Table 2:** Response to treatment and demographic variables.

| Variable | Category | Improved |  | No change or Worsened |  | p-value |
| --- | --- | --- | --- | --- | --- | --- |
|  |  | Count | Percent | Count | Percent |  |
| Gender | Female | 37 | 56.1 | 5 | 62.5 | 0.728 |
|  | Male | 29 | 43.9 | 3 | 37.5 |  |
| Age group | Youngest – 30 | 14 | 21.2 | 1 | 12.5 | 0.364 |
|  | 31-39 | 15 | 22.7 | 0 | 0.0 |  |
|  | 40-49 | 14 | 21.2 | 1 | 12.5 |  |
|  | 50-60 | 11 | 16.7 | 3 | 37.5 |  |
|  | 61-70 | 9 | 13.6 | 2 | 25.0 |  |
|  | 71-80 | 3 | 4.5 | 1 | 12.5 |  |
| Recoded Age | Youngest -49 | 43 | 65.2 | 2 | 25.0 | 0.028 |
|  | 50-Oldest | 23 | 34.8 | 6 | 75.0 |  |
| Race | African American | 3 | 4.7 | 0 | 0.0 | 0.374 |
|  | Asian | 1 | 1.6 | 1 | 12.5 |  |
|  | Caucasian | 45 | 70.3 | 5 | 62.5 |  |
|  | Multiple | 4 | 6.3 | 0 | 0.0 |  |
|  | Other | 11 | 17.2 | 2 | 25.0 |  |
| Ethnicity | Non-Hispanic | 40 | 60.6 | 6 | 75.0 | 0.428 |
|  | Hispanic | 26 | 39.4 | 2 | 25.0 |  |
| Residence | Apartment | 2 | 30 | 0 | 0.0 | 0.284 |
|  | Condo | 1 | 1.5 | 1 | 12.5 |  |
|  | Multigenerational | 3 | 4.5 | 0 | 0.0 |  |
|  | Single Family Home | 60 | 90.9 | 7 | 87.5 |  |
| Lives with | Alone | 3 | 4.8 | 1 | 12.5 | 0.678 |
|  | Spouse/Significant other | 54 | 85.7 | 7 | 87.5 |  |
|  | Parent | 4 | 6.3 | 0 | 0.0 |  |
|  | Other | 2 | 3.2 | 0 | 0.0 |  |

Table 3 presents the results for the response to treatment by smoking status and other drug use. No significant differences were found by smoking status, marijuana, vaping, or other intranasal drug use. No members of the sample reported IV drug use.

**Table 3:**
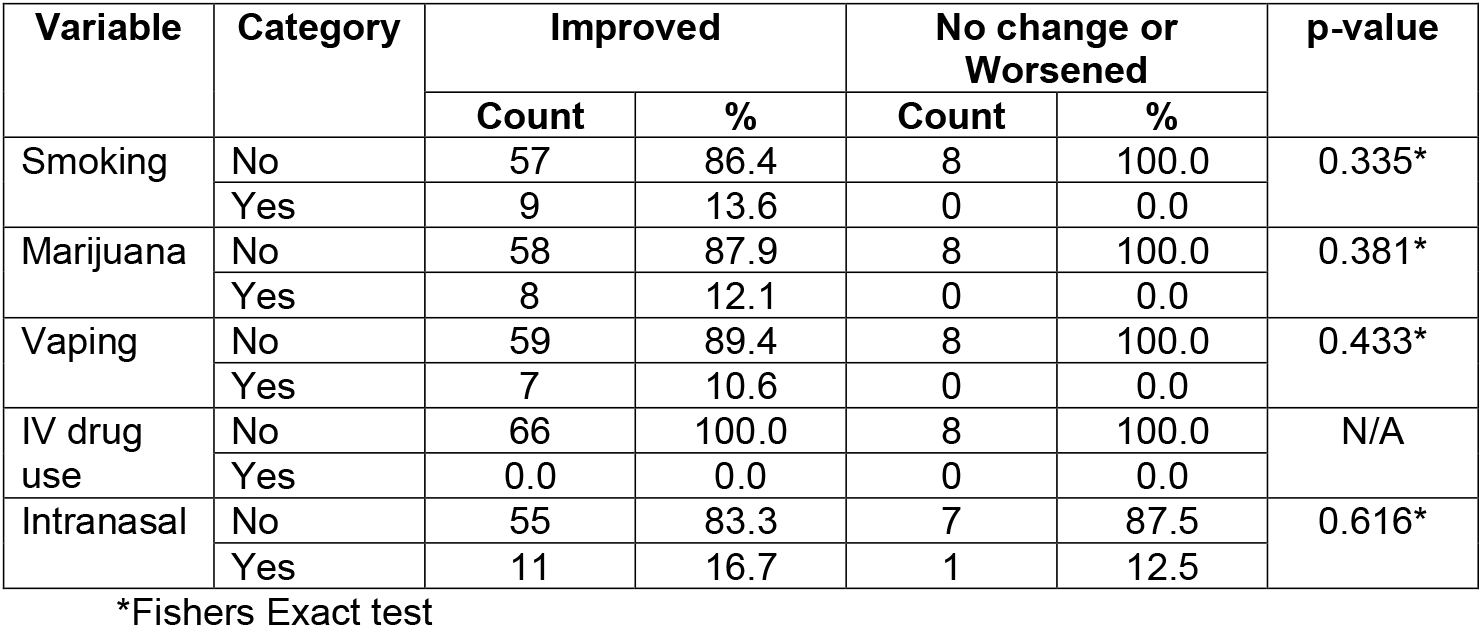
Response to treatment by smoking and drug use.

| Variable | Category | Improved |  | No change or Worsened |  | p-value |
| --- | --- | --- | --- | --- | --- | --- |
|  |  | Count | % | Count | % |  |
| Smoking | No | 57 | 86.4 | 8 | 100.0 | 0.335* |
|  | Yes | 9 | 13.6 | 0 | 0.0 |  |
| Marijuana | No | 58 | 87.9 | 8 | 100.0 | 0.381* |
|  | Yes | 8 | 12.1 | 0 | 0.0 |  |
| Vaping | No | 59 | 89.4 | 8 | 100.0 | 0.433* |
|  | Yes | 7 | 10.6 | 0 | 0.0 |  |
| IV drug use | No | 66 | 100.0 | 8 | 100.0 | N/A |
|  | Yes | 0.0 | 0.0 | 0 | 0.0 |  |
| Intranasal | No | 55 | 83.3 | 7 | 87.5 | 0.616* |
|  | Yes | 11 | 16.7 | 1 | 12.5 |  |

\*Fishers Exact test

Table 4 presents the results of treatment efficacy as related to other comorbid conditions. There were no statistical differences in any of the comorbid conditions as associated with treatment efficacy indicating that the treatment is well tolerated across all of these conditions that were collected.

**Table 4:**
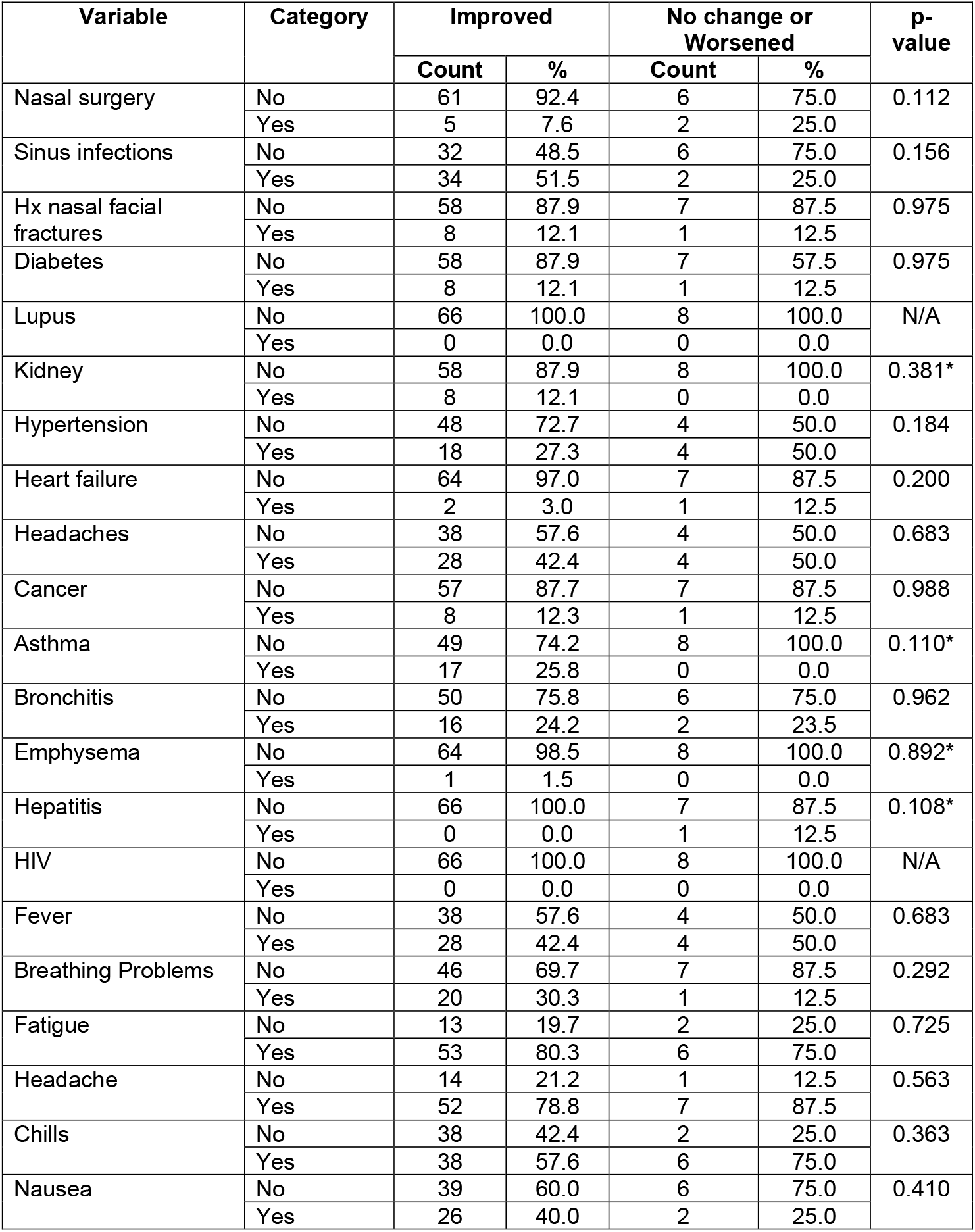
Response to treatment by comorbid conditions.

## DISCUSSION

The world has suffered from the coronavirus, COVID-19 pandemic. The primary mode of viral transmission has been the respiratory droplet, with the appreciation of the high affinity of the SARS-CoV-2 virus to the ACE 2 receptors of the nasal mucosa.^3^ Infected patients may continue to be carriers of the virus, and it is uncertain if the nasoepithelium serves as a potential reservoir of viral infection.^3, 7^ Moreover, patients infected with the virus may continue to pose a potential risk as evidence of viral shedding has been noted during asymptomatic infection as well as post recovery.^7^ An infected individual with coronavirus may be asymptomatic during the incubation period, or exhibit non-severe symptoms, or progress to life-threatening illness and death. Attempts to correlate clinical disease with SARS-CoV-2 viral loads from respiratory secretions, blood, and tissue have yielded conflicting results, as some patients with advanced disease have high viral loads while others do not.^2^ The use of nasal irrigation has been demonstrated to be effective at reducing symptoms of upper respiratory condition with hypertonic saline.^8^. The use of povidone iodine has been shown to reduce viral load in the Covid positive patient, however, in a small study, 42% of the participants experienced thyroid dysfunction during treatment.^9^ Nasal saline irrigations have been shown to be beneficial for COVID-19, however viral contamination of the rinse bottles and contact-induced transmission were of concern.^7^

Hypochlorous acid is an effective agent in vitro and in vivo. In vitro, the antiviral effect is dependent on the availability of the chloride ions, and its impact upon the viral nucleic acids DNA or RNA.^10,11^ In vivo, hypochlorous acid is naturally produced by the activated neutrophil during the oxidative burst phase by the enzyme myeloperoxidase. The activated neutrophil is estimated to produce 1.6 × 10-6 molecules of hypochlorous acid per second.^12^ Hypochlorous acid is FDA-approved as a preservative for saline solutions.^5^ Distinctive from other antiseptics such as povidone-iodine, ethanol, hydrogen peroxide, and chlorhexidine gluconate, hypochlorous acid cleansers are favorably non cytotoxic making them useful agents in wound care. Clinically, hypochlorous acid washes have been safely utilized in pediatric and adult wound care, involving skin, oral, and ocular usages.^4,5,6^ Its use in nasal lavage may potentially reduce viral load with the potential advantage of reducing the risk of surface contamination due to its antiseptic properties.

This study has demonstrated the safe utilization of hypochlorous acid solution (Vashe Wound Solution, Urgo Medical North America, LLC) for intranasal lavage in the ambulatory patient testing positive for the SARS-CoV-2 virus. Participants of this study demonstrated improvement in their clinical symptoms, and there were no statistical differences in any of the co-morbid conditions as associated with the intranasal lavage.

Mild adverse reactions include nasal burning sensation, mild self-limited epistaxis, and metallic taste associated with one participant with titanium dental implants. At 30-day follow up, no patients had required mechanical ventilation, and there were no deaths.

## LIMITATIONS

This study has several limitations: The sample size of the study was small. Although all patients were offered to participate, some declined to consider the option due to apprehensions associated with nasal rinsing. Because of the varied clinical presentations, the correlation of the start date of the nasal lavage with the presumed initial infection date was inconsistent. This was especially true for the patients who were asymptomatic or had very mild symptoms. In households with many occupants, it remains unclear if true isolation were achieved, especially in cases with young children or teenagers, both of whom were unable to participate due to the age-based inclusion criterion. No data was tabulated with regards to the number of participants who concurrently received outpatient monoclonal antibody therapy. No follow up nasal swabs or additional tests were performed after the nasal irrigation intervention.

Throughout the enrollment, no participants had received the COVID vaccine, and it remains uncertain how vaccination might impact clinical outcomes.

## CONCLUSIONS

This study does suggest the feasibility of using intranasal lavage with a hypochlorous acid solution for the COVID-19 positive ambulatory patient, however, further research is needed to appreciate its value and larger clinical strategy for prevention, relief, and treatment of disease. The potential effects of Vashe Wound Solution nasal lavage as a therapeutic agent in reducing viral load would need to be more thoroughly investigated as this is a novel application of this solution.

## Data Availability

All data produced in the present work are contained in the manuscript.

## Author Affiliation

TowerHealth Medical Group, Reading, PA

## Acknowledgement

The author acknowledges and thanks senior principal biostatistician Thomas E. Wasser PhD, MEd, CIM who performed the statistical data analysis. Additional thanks for the assistance of Dr. Ziad Osman, Medical Director of the TowerHealth Liggett Avenue Urgent Care Facility, and Alison Muller MLS (ASCP), MSPH, Trauma Research Coordinator.

